# How re-infections and newborns can change the visible and hidden epidemic dynamics?

**DOI:** 10.1101/2025.03.20.25324314

**Authors:** Igor Nesteruk

**Affiliations:** Institute of Hydromechanics, National Academy of Sciences of Ukraine, Kyiv, Ukraine; saac Newton Institute for Mathematical Sciences, University of Cambridge, UK

**Keywords:** mathematical modeling of infection diseases, SIR model, hidden epidemic dynamics, the COVIV-19 pandemic, pertussis epidemic in England

## Abstract

A recently proposed model for visible and hidden epidemic dynamics has been generalized to account for the effects of re-infections and newborns. The set of five differential equations and initial conditions contain 13 unknown parameters. The analysis of the equilibrium points, the examples of numerical solutions and comparisons with dynamics of real epidemics are provided. It was shown that equilibriums exist when the influence of re-infections or newborns can be neglected. A stable quasi-equilibrium for the particular case of almost completely hidden epidemics was revealed. Numerical results and comparisons with the COVID-19 epidemic dynamics in Austria and South Korea showed that re-infections, newborns and hidden cases make epidemics endless. The newborns can cause repeating epidemic waves even without re-infections. In particular, numerical simulations of the pertussis epidemic in England in 2023 and 2024 demonstrated that the next epidemic peak is expected in 2031. The proposed model can be recommended for calculations and predictions of visible and hidden numbers of cases, infectious and removed patients. With the use of effective algorithms for parameter identification, the accuracy of method could be rather high.

## 1. Introduction

Children born during an epidemic can significantly increase the number of susceptible patients, since many of them do not have immunity. To take into account this fact, a generalization the classical SIR (susceptible-infectious-removed) model [1-7] was proposed in [4]. Re-infections, typical for many infections including SARS-COV-2 [8-10], were simulated in [11]. Almost all infections have many asymptomatic and unregistered cases [12-18] influencing the epidemic dynamics [19-20]. In this study we propose the most general mathematical model which takes into account all the above factors. We will analyze the equilibrium points of corresponding set of ordinary differential equations. Some examples of numerical solutions reflecting the dynamics of the COVID-19 pandemic in South Korea and Austria and of the pertussis epidemic in England in 2023 and 2024 [21] will be presented.

## 2. Differential equations, initial conditions and parameter identification procedure

For every epidemic wave *i*, let us suppose the constant rate of increasing the numbers of susceptible persons *μ*_*i*_ (due to newborns) and divide the compartment of infectious persons *I(t)* (*t* is time) into visible (registered) and hidden (invisible/asymptomatic and unregistered) parts *I* = *I* ^(*v*)^ + *I* ^(*h*)^. As in [19, 20], we suppose that these persons are appearing according to the visibility coefficient *β*_*i*_ ≥ 1 and removing with rates 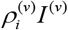and 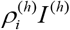, respectively. The compartment of removed persons *R(t)* is also divided into visible (registered) and hidden parts *R* = *R*^(*v*)^ + *R*^(*h*)^. As in [11], let us suppose the rates of re-infections to be proportional to the numbers of removed persons 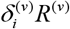 and 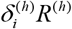. Then the general SIR model takes the following form:

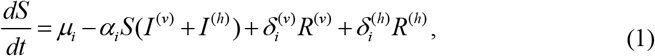

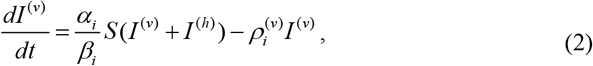

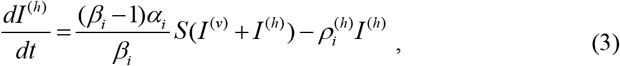

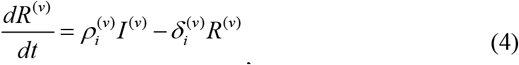

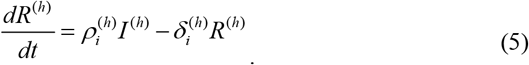

Infection, removal and re-infection rates 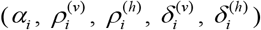, the visibility coefficient *β*_*i*_ and the increasing rate of the susceptible persons *μ*_*i*_ are supposed to be constant for every epidemic wave, i.e. for the time periods: 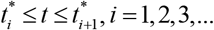.

Summarizing eqs. (1)–(5) yields non-zero value of the derivative:

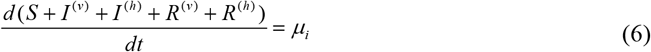

and the solution of the differential equation (6)

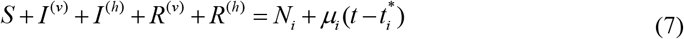

As in [7, 19, 20], we will consider the value *N*_*i*_ to be an unknown parameter of the model corresponding to the *i-th* wave, which is not equal to the known volume of population and must be estimated by observations. There is no need to assume that before the outbreak all people are susceptible (see, e.g., [4]), since many of them are protected by their immunity, distance, lockdowns, etc. Thus, we will not reduce the problem to a 4-dimensional one. It means that the solution can be obtained by numerical integration of the set of 5 differential equations (1)–(5) with the use of initial conditions:

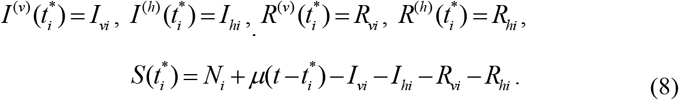

If at moment 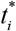 all previously infected persons are removed, we can take into account only cases starting to appear during *i-th* wave and use the following values of parameters:

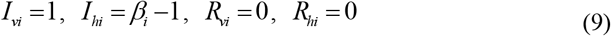

In comparison with the zero re-infections case [19], the accumulated numbers of registered cases *V*^*(v)*^ is no more equal to *I*^*(v)*^ + *R*^*(v)*^. We can state only that the derivative of this function (daily or monthly numbers of new cases) equals:

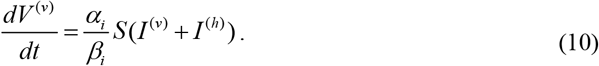

To determine *V*^*(v)*^ values, we have to integrate (10) as follows:

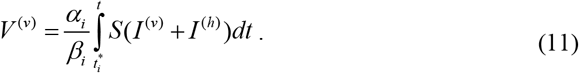

(the accumulation of cases has started at the moment 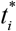).

The values of 13 parameters 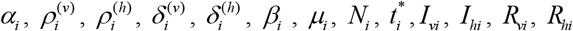 are unknown and have to determined based on the results of observations (e.g., accumulated numbers of visible cases 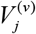 registered at moments of time *t, j* = 1, 2,…, *n*).

In particular, the method of least squares [22] can be used:

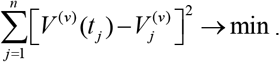

The values *V* ^(*v*)^ (*t*_*j*_) can be calculated using (11). To estimate the values of all parameters we need at least 13 observations. Since the results of observations are random, the accuracy of parameter identification increases at large numbers of *n*. On the other hand, a larger number of observations requires a longer time during which the parameters can change and cannot be considered to be constant. The experience of determining the optimal values of 4 parameters of the classical SIR model for the first waves of the COVID-19 pandemic showed that 14 observations are enough for fairly accurate and long-term forecasts [7]. Since the set of three differential equations corresponding to classical SIR model has an exact solution [7], the calculations of the optimal values of 4 parameters do not require a lot of time. The set of differential equations (1)–(5) can be solved only numerically and identification of 13 parameters need using the high performance computing, parallel codes or/and AI methods.

## 3. Equilibrium points

Let us find the values *S*^*^, *I*^*(*v*)^, *I*^*(*h*)^, *R*^*(*v*)^, *R*^*(*h*)^ corresponding to zero derivatives in the left-hand side of eqs. (1)–(5). These equilibrium points will give us the endemic characteristics of a disease. It follows from (2) and (3) that:

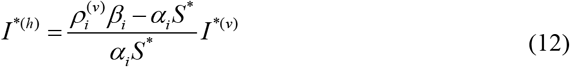

If *I*^*(*v*)^ ≠ 0, then eqs. (3) and (12) yield:

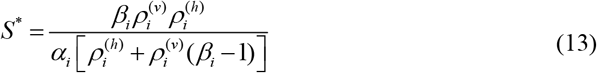

It follows from (1) and (12) that:

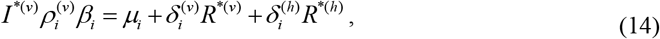

Eqs. (4), (5), (12), and (14) yield:

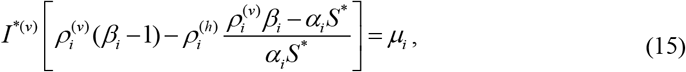

Taking into account (13) it can be shown that expression in square brackets equals zero.

Then non-trivial equilibrium (*I*^*(*v*)^ ≠ 0) occurs only at *μ*_*i*_ =0 with arbitrary values of *I*^*(*v*)^. Corresponding characteristics *I* ^*(*h*)^ and *S*^*^ can be calculated from (12), (13) and the equilibrium numbers of removed persons with the use of (4), (5) and (12):

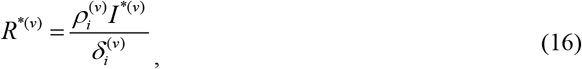

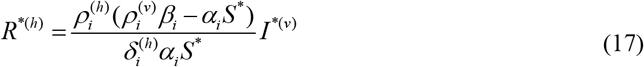

The endemic characteristics of fully visible epidemic (*β*_*i*_ =1) and the stability of the equilibrium were considered in [11].

When re-infections can be neglected (e.g., for pertussis), 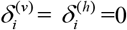 and eqs. (1)–(3) do not depend on eqs. (4) and (5). Eq. (12) holds for the equilibrium points of the set (1)–(3) and after its substituting into (1), we can obtain:

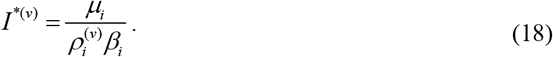

The analysis of the stability of this equilibrium can be performed numerically after the parameter identification. Here we consider particular cases, when this analysis can be done analytically.

If removing rates for visible and hidden patients are equal 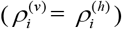,summarizing (2) and (3) yields the equation:

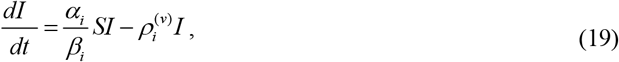

where *I* = *I* ^(*v*)^ + *I* ^(*h*)^ is the total number of infectious persons. Without re-infections, the set of equations (1), (19) does not depend on (3) and the equilibrium values are:

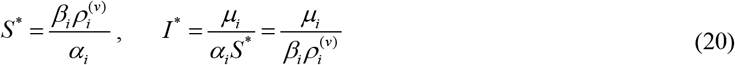

Jacobian matrix [23-25] for the set of differential eq. (1), (19) is equal to:

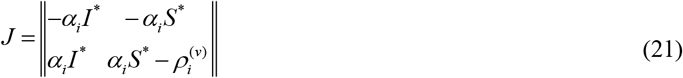

Taking into account (20), Jacobian (21) can be written as follows:

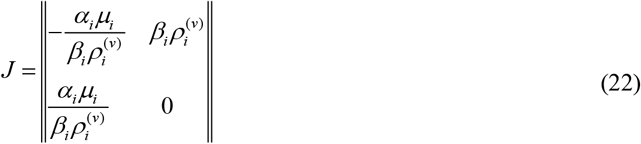

The eigenvalues of (22)

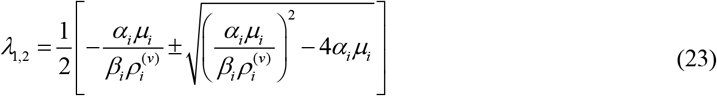

have negative real part, thus the equilibrium is stable [23-25]. The case of fully visible epidemic (*I* = *I* ^(*v*)^) can be easily obtained from (20)-(23) by putting *β*_*i*_=1.

It follows from (10), (20) that numbers of new cases increase at constant rate:

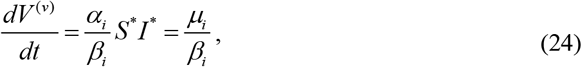

when this equilibrium is reached. According to (24), the total numbers of cases and susceptible persons increase at equal rate *μ*_*i*_. Pertussis in England demonstrated such endemic state in 2018 and 2019 with the average monthly numbers of new cases 246 =2948/12 and 307=3680/12, respectively (see [21] and Supplementary Fig. S1). Multiplying these values by the visibility coefficient, we can estimate the increase rate of susceptible newborns (see (24)). At moderate values of *β*_*i*_ this rate is much lower than the total birth rate (563,561 live births occurred in England in 2023, [26]). To decrease the endemic level, vaccinations of children and pregnant women are necessary, since at fixed birth rate only these interventions can decrease the value of parameter *μ*_*i*_.

## 4. Quasi-equilibrium states

Effective isolation of symptomatic patients can lead to the situation when *I* ^(*v*)^ ≪ *I* ^(*h*)^. Then it follows from (1) and (3), that

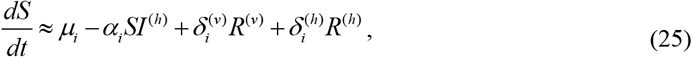

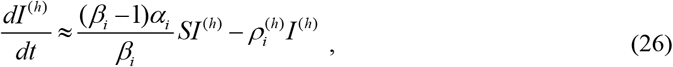

and a state with almost constant numbers of invisible infectious persons can occur at

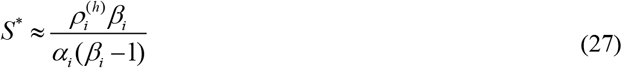

(if follows from (26) *dI* ^(*h*)^ / *dt* = 0). If numbers of removed persons are constant, eqs. at (4), (5) and (25) yield:

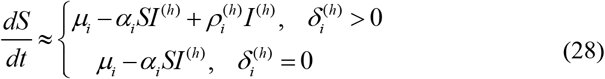

and approximately constant vales of susceptible persons at

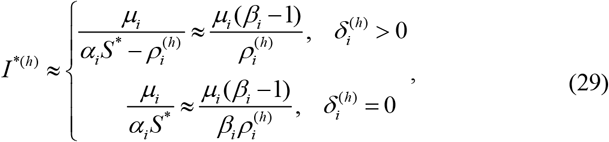

Jacobian matrix [23-25] for the set of approximate differential eq. (28) and (26) is equal to:

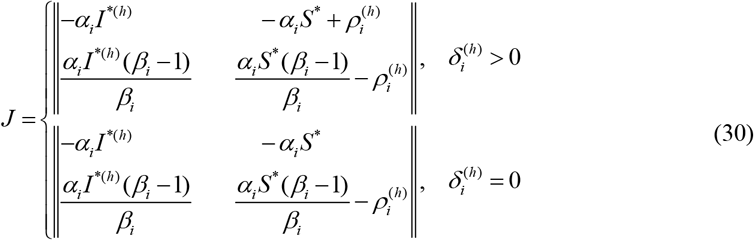

Taking into account (27), (29), Jacobian (30) can be written as follows:

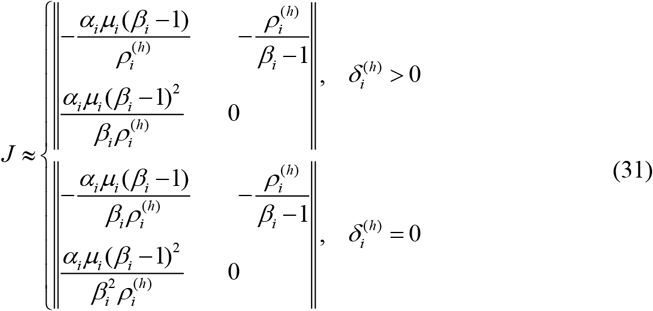

The eigenvalues of Jacobian (31)

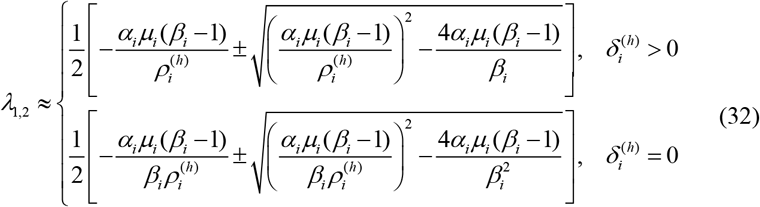

have negative real part at *β*_*i*_ >1, thus the quasi-equilibrium is stable [23-25] and it follows from (10), (27) and (29) that numbers of new visible cases increase at the approximately constant rate:

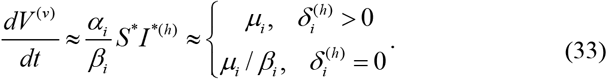

When re-infections occur 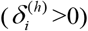 the increase rate of numbers of visible cases is approximately equal to *μ*_*i*_ and does not depend on infection, removal, re-infection, and visibility coefficients. Without re-infections, the numbers of visible cases increase at *β*_*i*_ times smaller rate (see (33)). In both cases the epidemic is driven by invisible (asymptomatic) infectious patients.

Probably such quasi-equilibrium state occurred in South Korea, Austria, Spain, and France in June 2020 between first and second waves of the COVID-19 pandemic (see average daily numbers of visible cases calculated in Chapter 8 of book [7]). For example, in South Korea in June 2020, the smoothed daily numbers of new cases *DC* varied from 38 to 48.1 (see black “crosses” in Fig. 1 representing dataset from [27], version uploaded on 23 December 2023) with the average value 44.4=(12799-11468)/30, which according to (33) could be equal to *μ*_*i*_. The relative variation is approximately 23% (10.1/44.4). In 2020 in South Korea 272,337 birth were registered, [28] (the average value is 744 per day). Thus, only 6% of newborns become susceptible.

**Fig 1.**
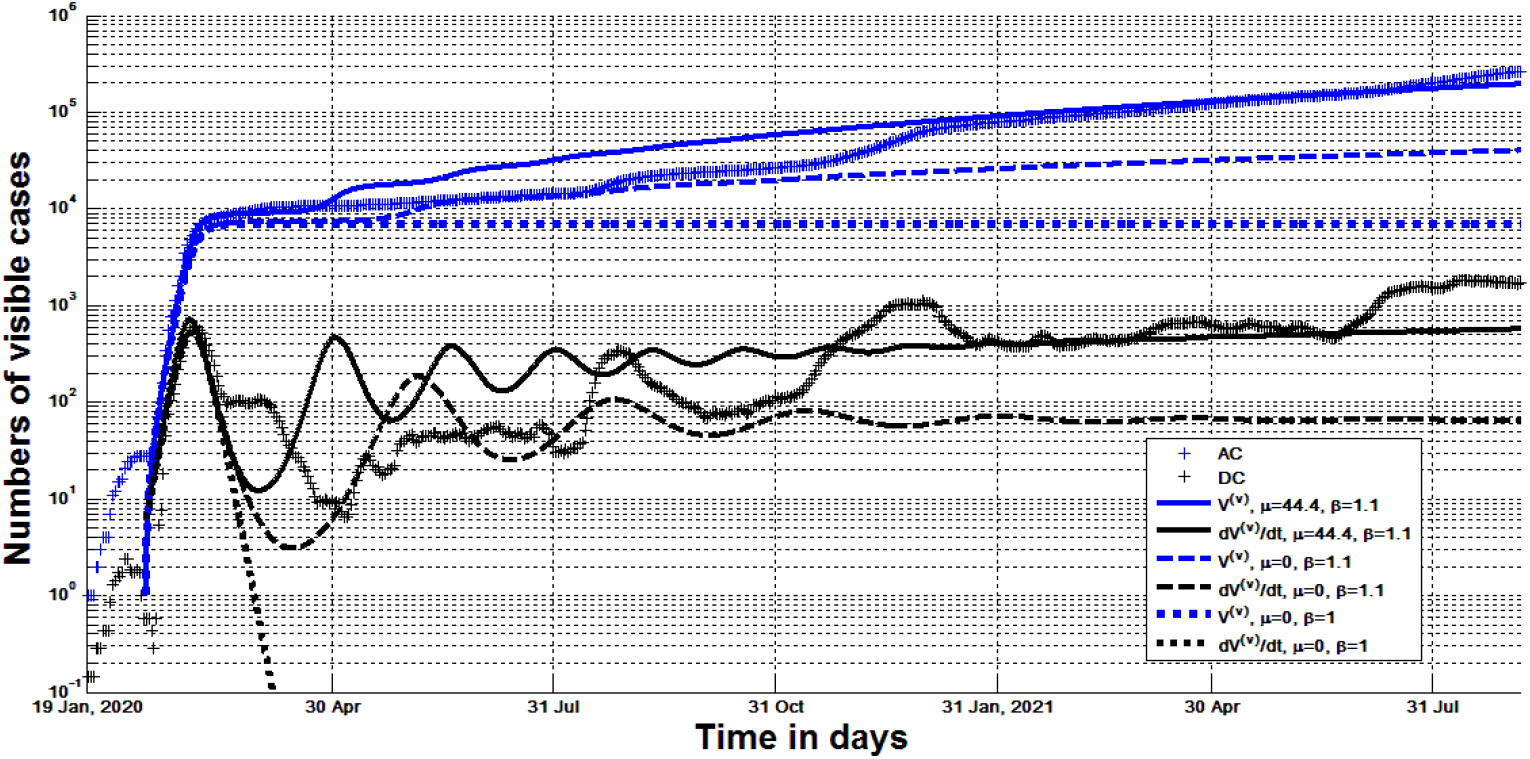
Visible numbers of COVID-19 cases in South Korea in 2020 and 2021. Blue and black “crosses” represent visible (registered) accumulated numbers *AC* and smoothed daily numbers of new cases *DC*, respectively, [27]. Curves show the results of numerical integration of (1)-(5) at different values of parameters: blue - *V* ^(*v*)^; black - *dV* ^(*v*)^ / *dt*.

In Austria, the smoothed daily numbers of new cases varied from 24.6 to 35.4 in the period from 27 May to 24 June, 2020 (see black “crosses” in Fig. 2 representing dataset from [27], version of 23 December 2023) with the average value 31.1, and relative variation 36%. The average daily number of newborns was 243 in 2020 (the figure was calculated with the use of [28, 29]), 12.8% of which become susceptible. Comparison of percentage of susceptible newborns allows us to conclude that in June 2020 tracing and isolation of infectious in South Korea were more effective. It must be noted that many countries (e.g., Ukraine, the UK, the US, Italy, Germany, Sweden, Republic of Moldova) have not achieved the quasi-steady state and in some of them, the second COVID-19 wave started before finishing the first one, [7]. Probably, in these countries, the efficacy of isolation of infectious patients was not enough to have *I* ^(*v*)^ ≪ *I* ^(*h*)^.

**Fig 2.**
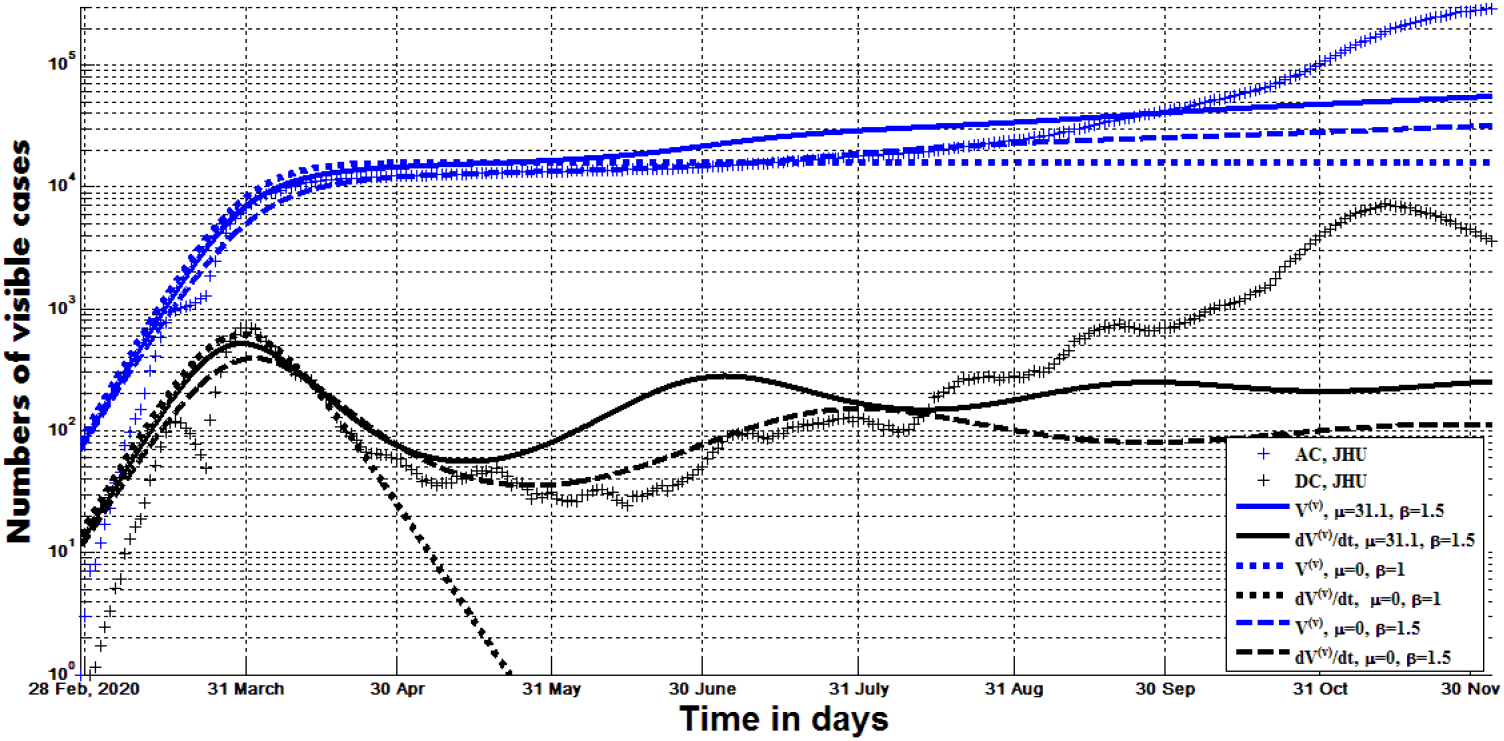
Visible numbers of COVID-19 cases in Austria in 2020. Blue and black “crosses” represent visible (registered) accumulated numbers *AC* and smoothed daily numbers of new cases *DC*, respectively, [27]. Curves show the results of numerical integration of (1)-(5) at different values of parameters: blue - *V* ^(*v*)^; black - *dV* ^(*v*)^ / *dt*.

Existence of the non-trivial stable quasi-equilibrium, makes epidemics endless, since newborns are in every country and it is very difficult to isolate all asymptomatic infectious persons. Moreover, in the case of complex eigenvalues (32) the epidemic waves can repeat with the period:

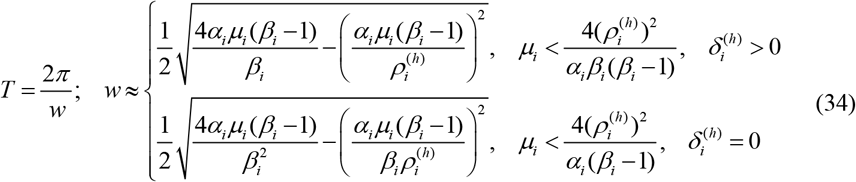

since in the vicinity of equilibrium point the linearized set of differential equations has oscillatory solutions [4, 25]. For example, in 2020 and 2021, Zero-COVID countries [31] were trying to stop community transmission of SARS-Cov-2 infection with the use of contact tracing, mass testing, border quarantine, lockdowns, and mitigation software. In particular, in Hong Kong the smoothed daily numbers of new COVID-19 cases were less than 20 per million in 2020 and 2021, [27], i.e., the epidemic was controlled but not removed completely, since in 2022 a huge epidemic wave occurred [27]. Other Zero-COVID countries have also experienced severe pandemic waves after reducing quarantine limitations and decreasing the test per case ratio [32].

Similar oscillatory solution can exist for the stable equilibrium discussed in the previous Section 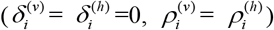. It follows from (23) that corresponding frequency equals:

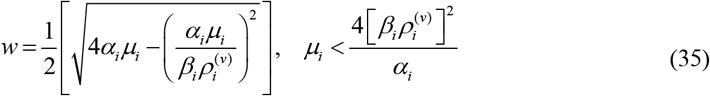

## 4. Examples of numerical solutions

The set of differential equations (1)–(5) were integrated numerically with the use of initial conditions (8), (9) and the fourth order Runge-Kutta method [25]. The results were tested with the use of exact solutions for the classical SIR model [7, 19, 33] and shown in Figs. 1-6 for different values of parameters. Blue curves represent the accumulated numbers of visible cases *V* ^(*v*)^; black ones – the theoretical estimations of the daily numbers of visible cases *dV* ^(*v*)^ / *dt*. The daily numbers of hidden cases have the same trends, since according to (3), (10) 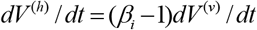.No method of least squares was used to detect the optimal values of parameters. We have used only parameter sets that provide some similarity to the observed numbers of accumulated (*AC*) and daily cases (*DC*) registered after outbreaks and shown by blue and black “crosses”, respectively.

Figs. 2 and 3 illustrate the COVID-19 pandemic dynamics in Austria. Solid lines correspond to the values of parameters: 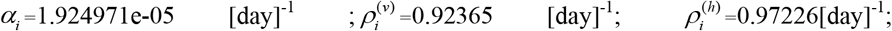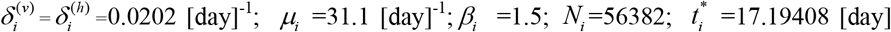 (zero value of time corresponds to 15 January 2020); *I*_*vi*_ =1; *I*_*hi*_ =0.5; *R*_*vi*_ = *R*_*hi*_ =0. The dashed curves correspond the same values of parameters, but zero birth rate *μ*_*i*_ =0. Dotted lines correspond to the fully visible epidemic with 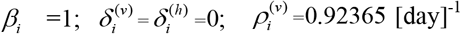, which can be simulated with the use of classical SIR model [7]. The optimal values of parameters calculated in [7] based on a previous versions of *AC* dataset [27] differ from the listed above set of classical SIR model parameters.

**Fig 3.**
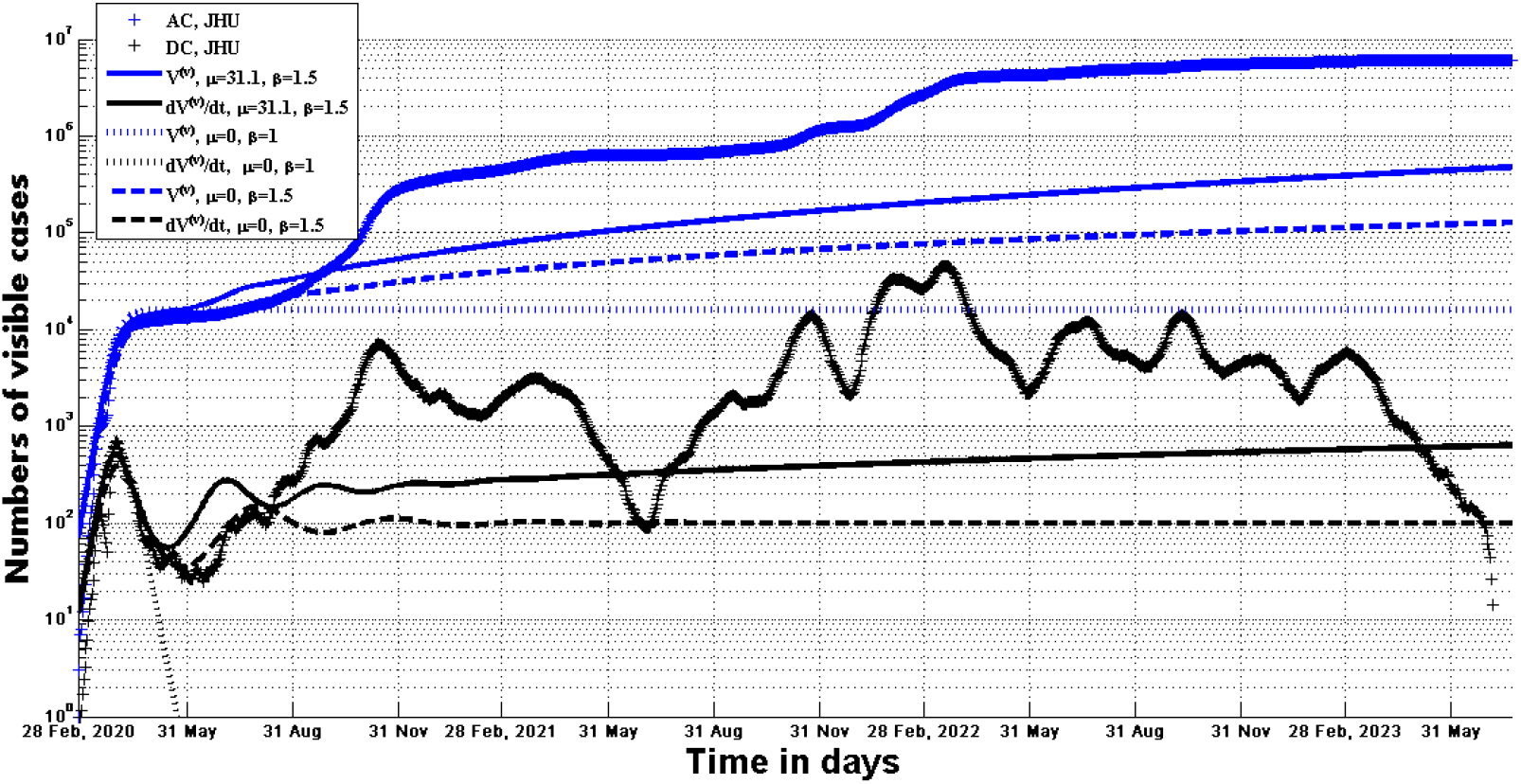
The COVID-19 pandemic dynamics in Austria. Visible accumulated (blue) and daily (black) cases. Blue and black “crosses” represent visible (registered) accumulated numbers *AC* and smoothed daily numbers of new cases *DC*, respectively, [27]. Curves show the results of numerical integration of (1)-(5) at different values of parameters: blue - *V* ^(*v*)^; black - *dV* ^(*v*)^ / *dt*.

Figs. 1 and 4 illustrate the COVID-19 pandemic dynamics in South Korea. Solid lines correspond to the values of parameters:

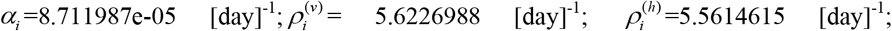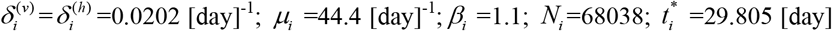 (zero value of time corresponds to 15 January 2020); *I*_*vi*_ =1; *I*_*hi*_ =0.5; *R*_*vi*_ = *R*_*hi*_ =0. The dashed curves correspond the same values of parameters, but zero birth rate *μ*_*i*_ =0. Dotted lines correspond to the fully visible epidemic with 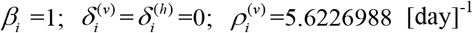, which can be simulated with the use of classical SIR model [7]. The optimal values of parameters calculated in [7] based on a previous versions of *AC* dataset [27] differ from the listed above set of classical SIR model parameters.

**Fig 4.**
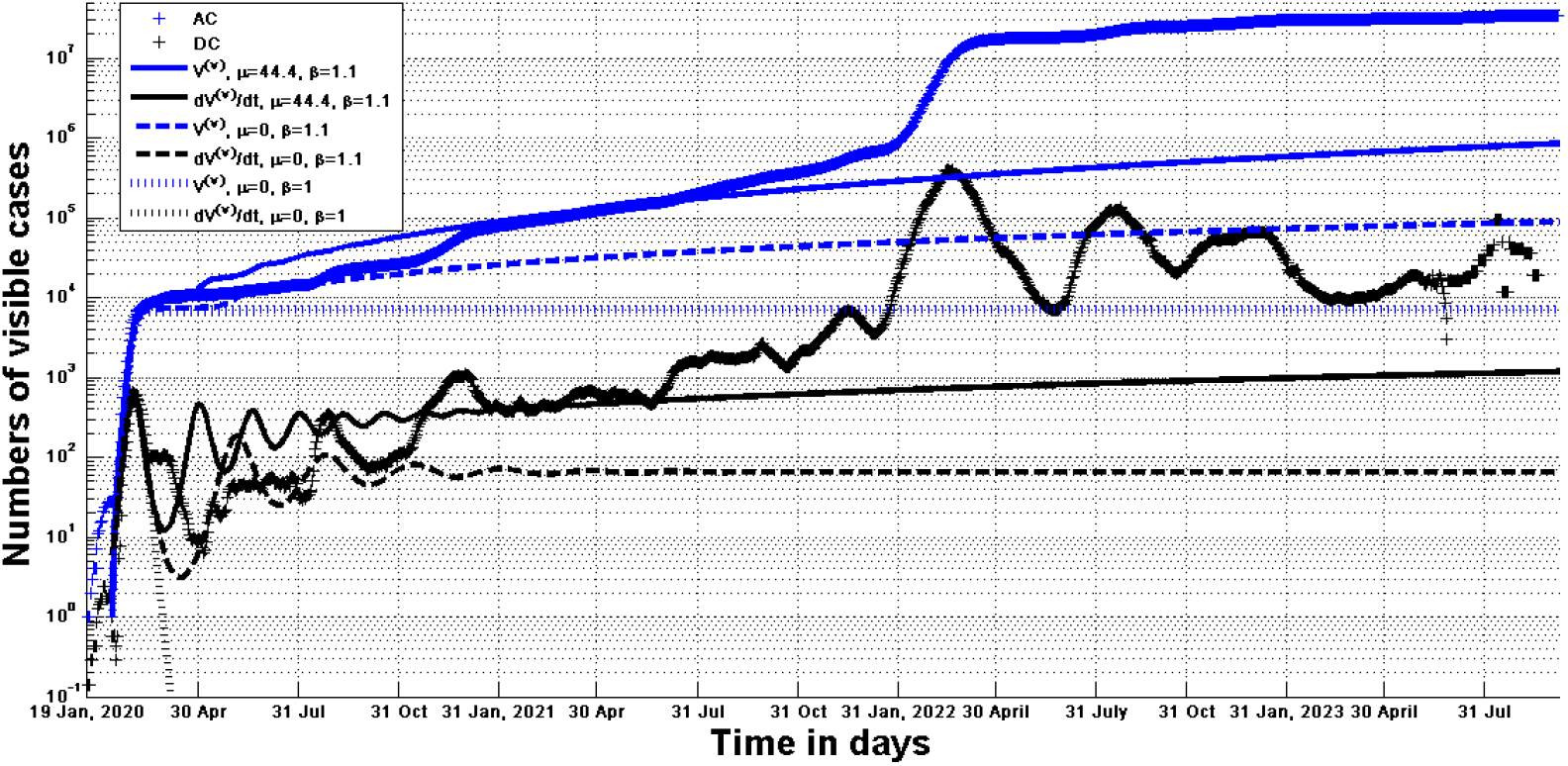
The COVID-19 pandemic dynamics South Korea. Visible accumulated (blue) and daily (black) cases. Blue and black “crosses” represent visible (registered) accumulated numbers *AC* and smoothed daily numbers of new cases *DC*, respectively, [27]. Curves show the results of numerical integration of (1)-(5) at different values of parameters: blue - *V* ^(*v*)^; black - *dV* ^(*v*)^ / *dt*.

The classical SIR model yields very optimistic predictions of the epidemic duration and final numbers of visible cases (see dotted curves in Figs. 1-4). Re-infections yield infinite epidemic durations even at zero birth rate 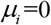 (see dashed curves in Figs. 1-4 which tend to a non-trivial equilibrium *I*^*(*v*)^ ≠ 0). Corresponding characteristics *I* ^*(*h*)^, *S*^*^, *R*^*(*v*)^ and *R*^*(*h*)^ can be calculated with the use of (12), (13), (16), and (17). Then the constant equilibrium numbers of new daily visible cases *DC*^*\**^ can be determined from eq. (10) as follows:

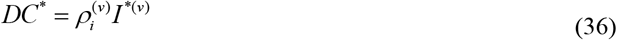

South Korea has stopped to report new COVID-19 cases at rather high level of *DC* (see black “crosses” in Fig. 4). Eq. (36) demonstrates that the numbers of infectious persons in South Korea remained high in August 2023. In Austria, there was a decreasing *DC* trend before stop the reporting in June 2023 (see black “crosses” in Fig. 3).

Solid curves in Figs. 1-4 represent the case *μ*_*i*_ > 0 and show no equilibrium tends (as mentioned in Section 3). Both *V* ^(*v*)^ (blue) and *dV* ^(*v*)^ / *dt* (black) values increased in 2023 (see Figs. 3 and 4). The accumulated numbers of cases *V* ^(*v*)^ were quite consistent with *AC* values registered in South Korea before October 2021 (compare solid blue curves and “crosses” in Figs. 1 and 4). *AC* and *DC* numbers registered in Austria from mid-March to mid-August 2020 are rather close to the values *V* ^(*v*)^ and *dV* ^(*v*)^ / *dt* corresponding the case *μ*_*i*_ =0 (compare “crosses” and dashed curves in Fig. 2).

Re-infections cause oscillations in daily numbers of new cases (see solid and dashed black curves in Figs. 1-4) and create the illusion of different pandemic waves, although the model parameters were considered fixed throughout the time period shown in Figs. 1-4. To have better coincidence with the real dynamics (influenced by changing in quarantine restrictions, social behavior, new virus strains, vaccinations, etc.), simulations have to be repeated with the use of observations for different periods (for example, for February-March 2022 in South Korea, see Fig. 4).

Figs. 5 and 6 illustrate the dynamics of pertussis epidemic in England in 2023 and 2024. The accumulated registered numbers of cases are listed in Table S1 according to the dataset [21] (version available on 13 February 2025) and shown by blue “crosses”. The average daily numbers of new cases *DC* were calculated with the use of the formula presented in [33], are listed in Table S1 and shown by black “crosses”. Solid lines correspond to the values of parameters:

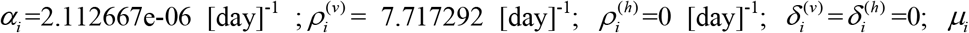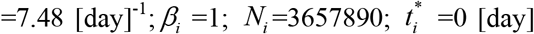 (zero value of time corresponds to 31 December 2022); *I*_*vi*_ =0.029115; *I*_*hi*_ = *R*_*vi*_ = *R*_*hi*_ =0. The dotted and dashed curves represent SIR simulations of the first and second pertussis waves, respectively (*μ*_*i*_ =0, *β*_*i*_ =1, the values of other parameters are listed in [19]). All simulations suppose that the epidemic is completely visible, i.e. *I* = *I* ^(*v*)^; *V* =*V* ^(*v*)^; *β*_*i?*_=1.

**Fig 5.**
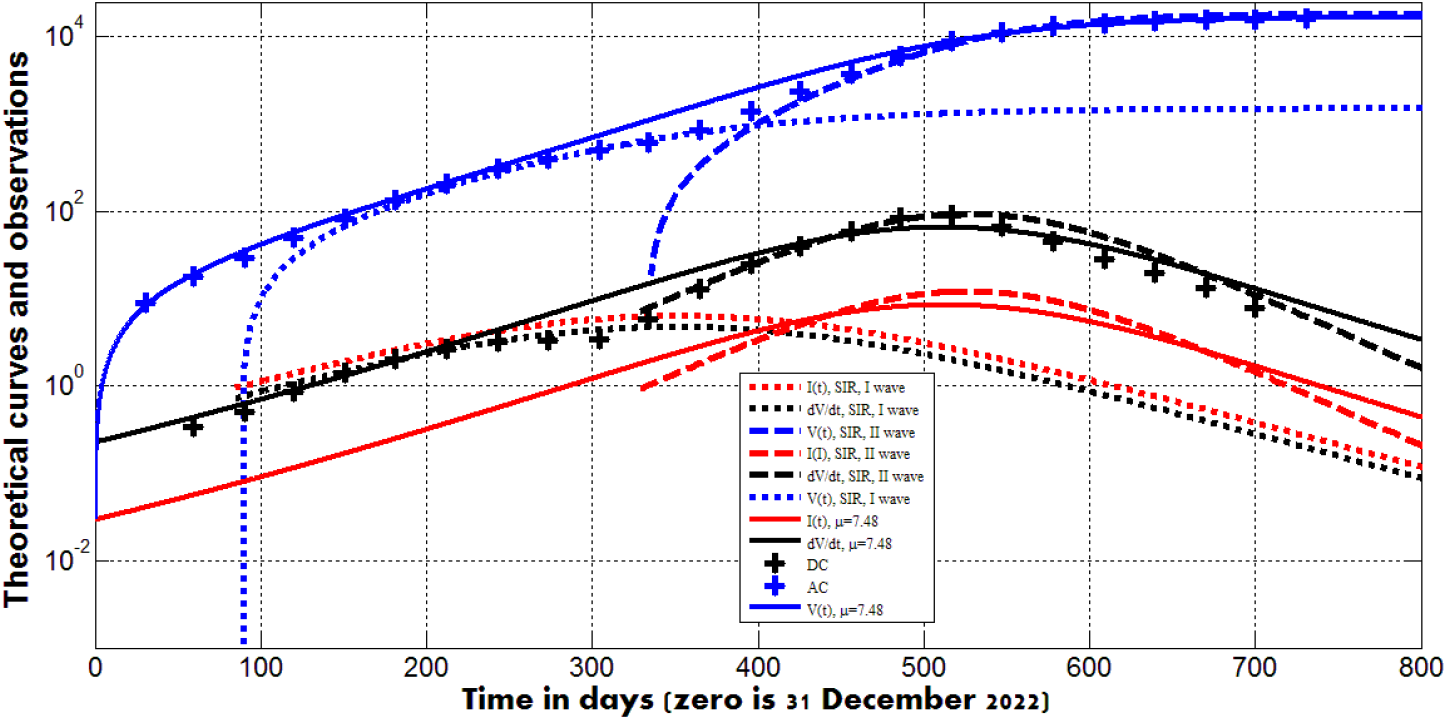
Dynamics of pertussis epidemic in England in 2023 and 2024. Accumulated cases (blue), average daily cases (black) and numbers of infectious persons (red). Blue and black “crosses” represent accumulated numbers *AC* and smoothed daily numbers of new cases *DC*, respectively, Table S1. Curves show the results of numerical integration of (1)- (5) at *β*_*i*_ =1: blue – *V* = *V* ^(*v*)^; black - *dV* / *dt*; red - *I* = *I* ^(*v*)^.

**Fig 6.**
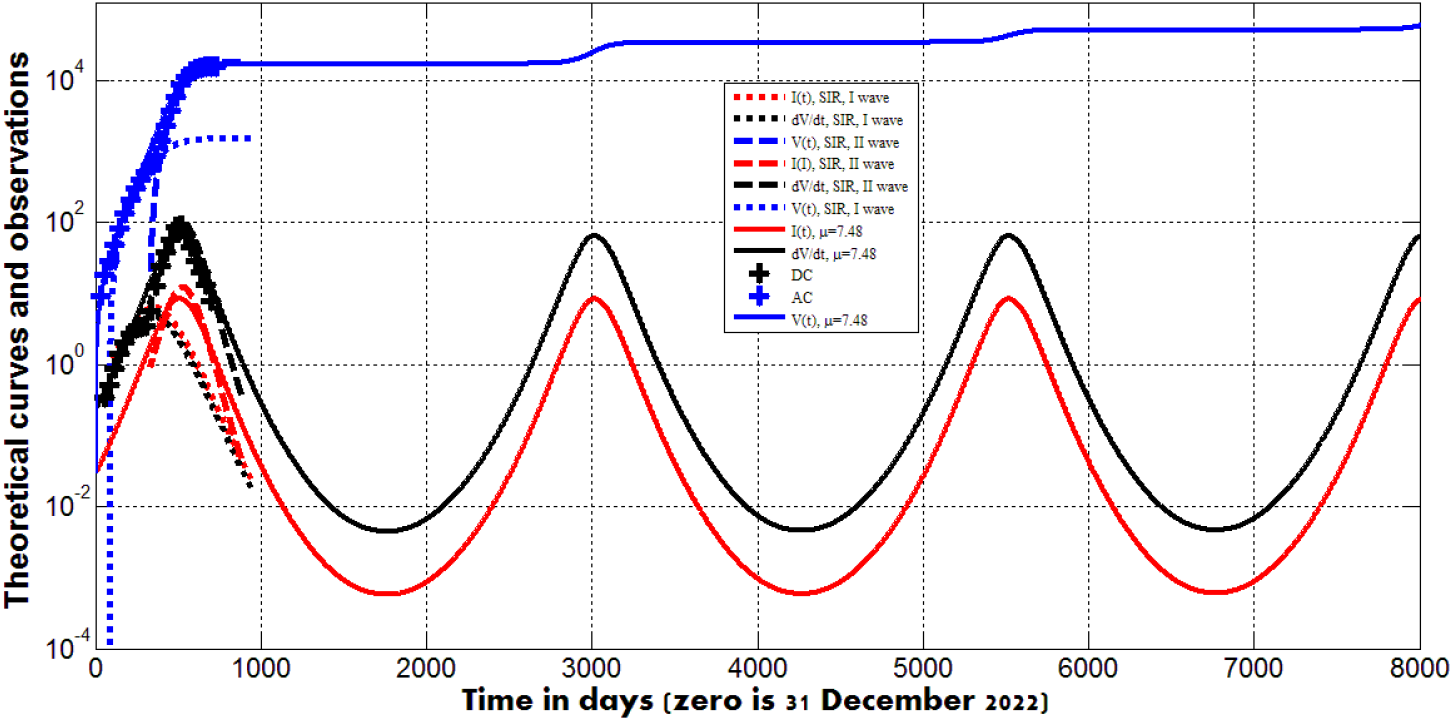
Predicted waves of pertussis epidemic in England. Accumulated cases (blue), average daily cases (black)and numbers of infectious persons (red). Blue and black “crosses” represent accumulated numbers *AC* and smoothed daily numbers of new cases *DC*, respectively, Table S1. Curves show the results of numerical integration of (1)- (5) at *β*_*i*_ =1: blue – *V* = *V* ^(*v*)^; black - *dV* / *dt*; red - *I* = *I* ^(*v*)^.

In comparison with SIR simulations at *μ*_*i*_ =0 (dotted and dashed curves), taking into account the birth rate (*μ*_*i*_ =7.48 [day]^-1^, solid curves) allowed us to obtain rather good agreement between the theory and results of all observations (compare blue and black solid curves with blue and black “crosses” in Fig. 5). Moreover, long epidemic waves with the repeating after around 2500 days (6.85 years) were revealed (see Fig. 6). Formula (35) yields the period of 1678 days (4.6 years) for the solution of linearized differential equations in the vicinity of the asymptotically stable spiral point [25] corresponding to eigenvalues (23). The difference can be explained by the large deviation of the numerical solution (red and black curves in Fig. 6) from the equilibrium values 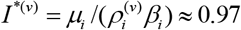 (see (18)) and 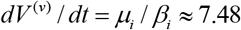 (see (24)). To decrease the sizes of new waves (maximum total daily numbers of new cases), the parameter *μ*_*i*_ has to be diminished (due to vaccinations of children and pregnant women).

According to [21], “pertussis is a cyclical disease that peaks every 3 to 5 years, with the last cyclical increase occurring in 2016 and the last major outbreak occurring in 2012”. The numbers of laboratory confirmed pertussis cases in England by quarter demonstrate that the previous peak was approximately 7.75 years before the peak registered in second quarter of 2024 (see Fig. S2, taken from [21]). Thus, the results of observations correlate with the theoretical findings, especially after taking into account the fact that several years is rather long time to use constant values of parameters. The accuracy of predictions can be improved with the use of optimal values of parameters. The application of the least squares method could be a topic of further investigations.

## 5. Conclusions

A recently proposed model for visible and hidden epidemic dynamics has been generalized to take into account the impact of re-infections and newborns. The set of five differential equations and initial conditions contain 13 unknown parameters, which makes it difficult to identify them using observational data. However, the analysis of the equilibrium points and the examples of numerical solutions provided allowed drawing important conclusions regarding the influence of various factors on the epidemic dynamics.

It was shown that in general case no equilibrium exists, but stable equilibriums are possible, when the influence of re-infections or newborns can be neglected. If the numbers of visible infectious are much lower then hidden ones, a stable quasi-equilibrium exists with constant daily numbers of new visible cases. Re-infections, newborns and hidden cases make epidemics endless. These facts were illustrated by numerical results and comparisons with the COVID-19 epidemic dynamics in Austria and South Korea. When re-infections can be neglected (e.g. for pertussis), the newborns can cause repeating epidemic waves. In particular, numerical simulations of the pertussis epidemic in England in 2023 and 2024 demonstrated that the next epidemic peak is expected in 2031.

The proposed model can be recommended for calculations and predictions of visible and hidden numbers of cases, infectious and removed patients. With the use of effective algorithms for parameter identification, the accuracy of method could be rather high.

## Clarification point

No humans or human data was used during this study

## Data availability

All data generated or analyzed during this study are included in this text.

## Acknowledgements

The author is grateful to Ulrike Tillmann, James Robinson, Robin Thompson, Matt Keeling, Paul Brown, and Oleksii Rodionov for their support and providing very useful information. This paper was written with the support of the INI-LMS Solidarity Programme at the University of Warwick, UK.

## Supplementary

**Fig S1.**
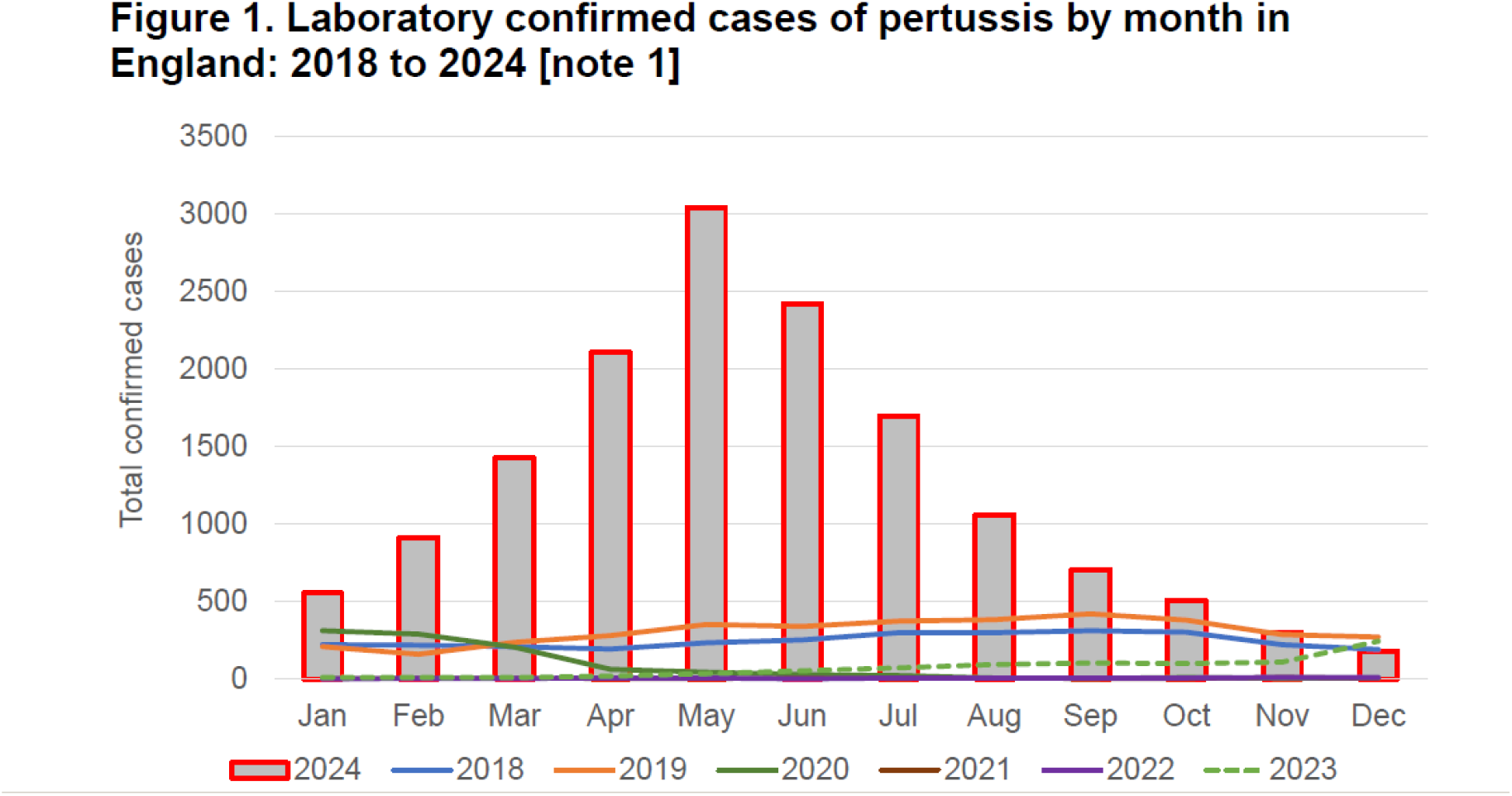
Monthly numbers of visible pertussis cases in England, [21].

**Fig S2.**
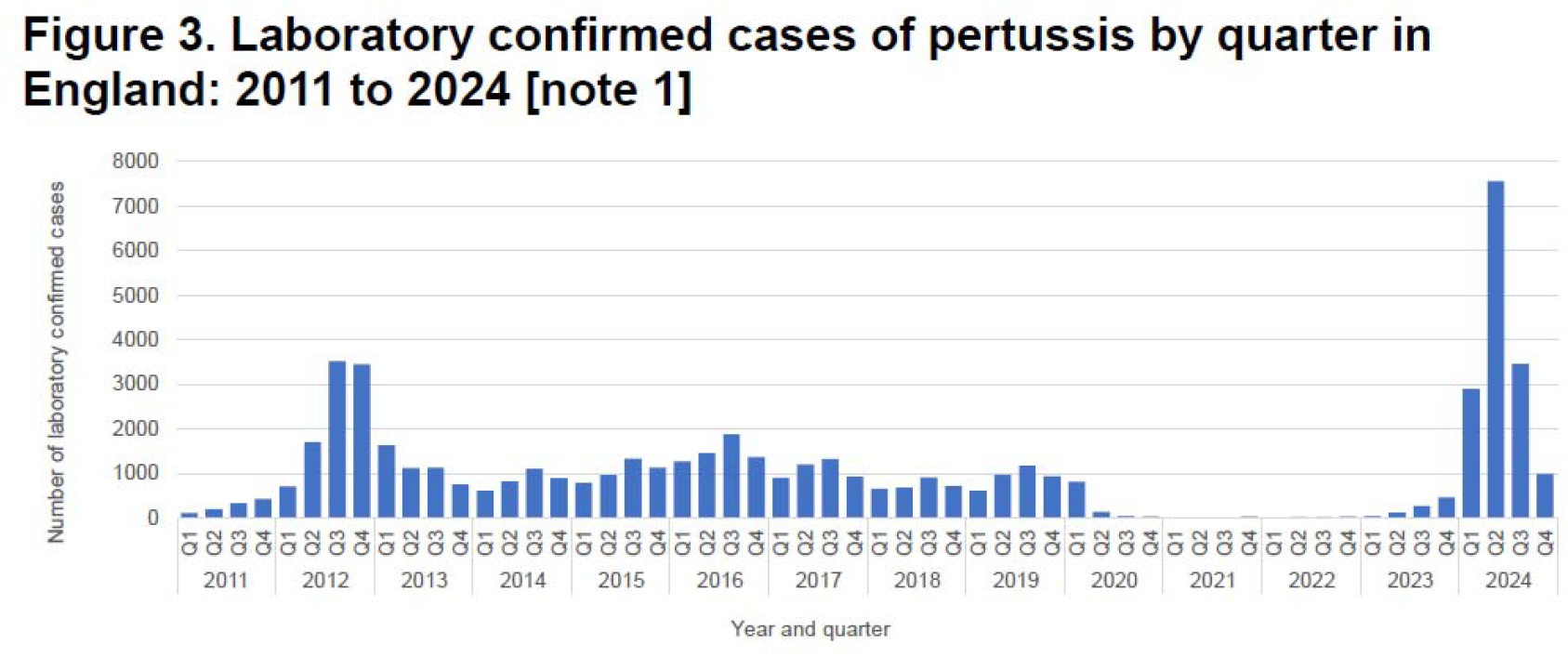
Numbers of visible pertussis cases by quarter in England, [21].

**Table S1.**
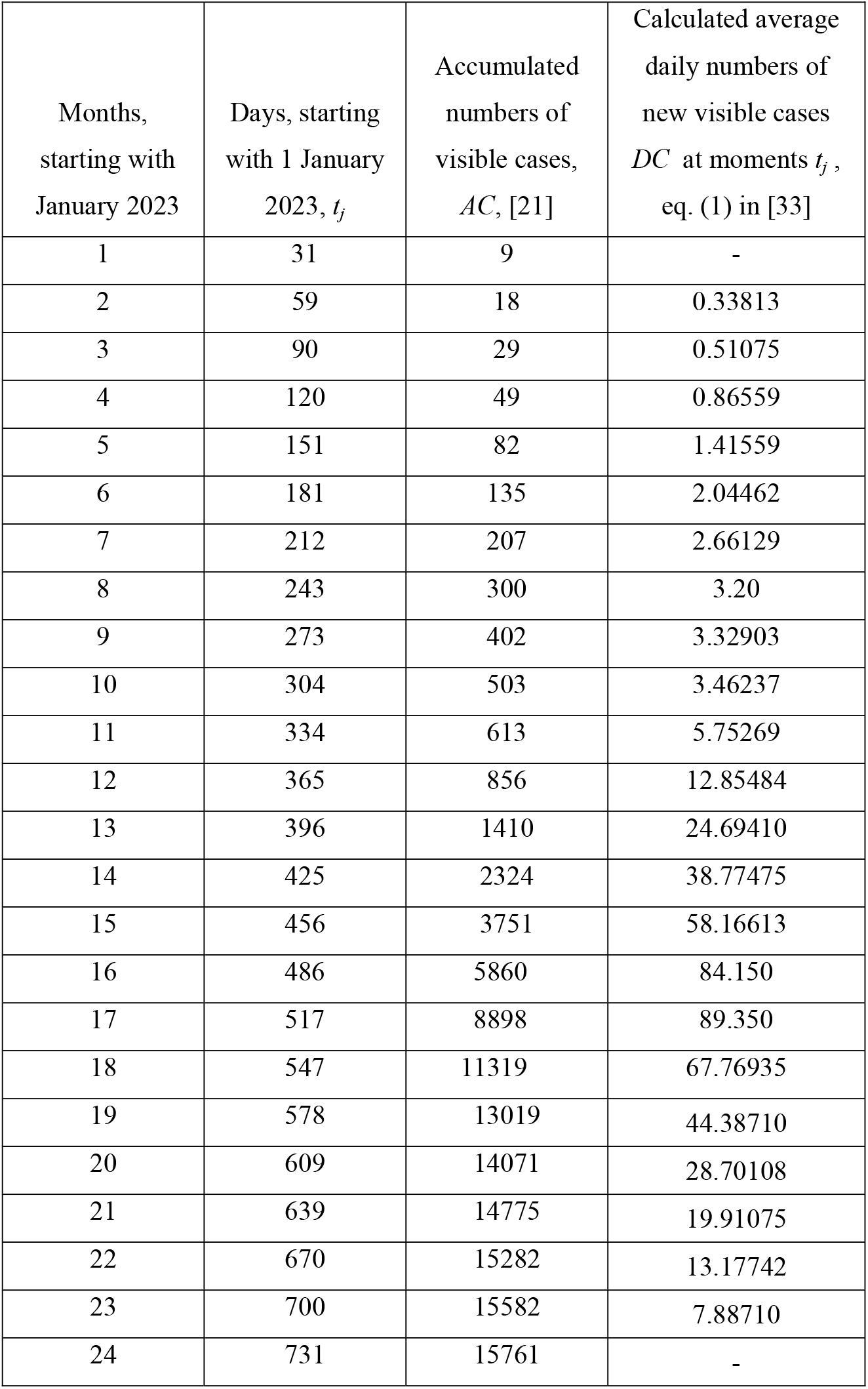
Accumulated numbers of confirmed pertussis cases in England in 2023 and 2024 (according to [21], version 13 February, 2025) and estimations of the average daily numbers of visible cases.

| Months,<br>starting with<br>January 2023 | Days, starting<br>with 1 January<br>2023, $t_j$ | Accumulated<br>numbers of<br>visible cases,<br>$AC$, [21] | Calculated average<br>daily numbers of<br>new visible cases<br>$DC$ at moments $t_j$,<br>eq. (1) in [33] |
| --- | --- | --- | --- |
| 1 | 31 | 9 | - |
| 2 | 59 | 18 | 0.33813 |
| 3 | 90 | 29 | 0.51075 |
| 4 | 120 | 49 | 0.86559 |
| 5 | 151 | 82 | 1.41559 |
| 6 | 181 | 135 | 2.04462 |
| 7 | 212 | 207 | 2.66129 |
| 8 | 243 | 300 | 3.20 |
| 9 | 273 | 402 | 3.32903 |
| 10 | 304 | 503 | 3.46237 |
| 11 | 334 | 613 | 5.75269 |
| 12 | 365 | 856 | 12.85484 |
| 13 | 396 | 1410 | 24.69410 |
| 14 | 425 | 2324 | 38.77475 |
| 15 | 456 | 3751 | 58.16613 |
| 16 | 486 | 5860 | 84.150 |
| 17 | 517 | 8898 | 89.350 |
| 18 | 547 | 11319 | 67.76935 |
| 19 | 578 | 13019 | 44.38710 |
| 20 | 609 | 14071 | 28.70108 |
| 21 | 639 | 14775 | 19.91075 |
| 22 | 670 | 15282 | 13.17742 |
| 23 | 700 | 15582 | 7.88710 |
| 24 | 731 | 15761 | - |

